# Disease correlates of quantitative susceptibility mapping rim lesions in multiple sclerosis

**DOI:** 10.1101/2021.05.29.21257734

**Authors:** Melanie Marcille, Sandra Hurtado Rúa, Charles Tyshkov, Abhishek Jaywant, Joseph Comunale, Ulrike W. Kaunzner, Nancy Nealon, Jai S. Perumal, Lily Zexter, Nicole Zinger, Olivia Bruvik, Yi Wang, Elizabeth Sweeney, Amy Kuceyeski, Thanh D. Nguyen, Susan A. Gauthier

**Affiliations:** Department of Neurology, Weill Cornell Medicine, New York, NY, USA; Department of Mathematics and Statistics, Cleveland State University, Cleveland, OH, USA; Department of Pediatrics, Weill Cornell Medicine, New York, NY, USA; Department of Psychiatry and Rehabilitation Medicine, Weill Cornell Medicine, New York, NY, USA; Department of Radiology, Weil Cornell Medicine, New York, NY, USA; Feil Family Brain and Mind Institute, Weill Cornell Medicine, New York, NY USA; Department of Population Health Sciences, Weill Cornell Medicine, New York, NY, USA

**Keywords:** multiple sclerosis, chronic active lesion, quantitative susceptibility mapping, disability, neurodegeneration

## Abstract

**Objective:** This study aimed to explore the association between chronic active rim+ lesions, identified as having a hyperintense rim on quantitative susceptibility mapping (QSM), on both clinical disability and imaging measures of neurodegeneration in patients with multiple sclerosis.

**Methods:** The patient cohort was composed of 159 relapsing remitting multiple sclerosis patients aged 42.17 ± 10.25 years and disease duration of 10.74 ± 7.51 years. The *Brief International Cognitive Assessment for Multiple Sclerosis* and Expanded Disability Status Scale (EDSS) were used to assess clinical disability. Cortical thickness and thalamic volume were evaluated as imaging measures of neurodegeneration.

**Results:** A total of 4,469 multiple sclerosis lesions were identified, of which 171 QSM rim+ (3.8%) lesions were identified among 57 patients (35.9%). In a multivariate regression model, as the overall total lesion burden increased, patients with at least one rim+ lesion on QSM performed worse on both physical disability and cognitive assessments, specifically the Symbol Digit Modalities Test (p=0.010), California Verbal Learning Test-II (p=0.030), and EDSS (p=0.001). In a separate univariate regression model, controlling for age (p<0.001), having at least one rim+ lesion was related to more cortical thinning (p= 0.03) in younger patients (< 45 years). Lower thalamic volume was associated with older patients (p=0.038) and larger total lesion burden (p<0.001) however, the association did not remain significant with rim+ lesions (p=0.10).

**Interpretation:** Our findings demonstrate the significant impact of chronic active lesions, as identified on QSM, on both clinical disability and imaging measures of neurodegeneration in patients with multiple sclerosis.

## Introduction

Innate immunity plays a pivotal role in the pathophysiology of multiple sclerosis (MS), and important cell types involved in this process are CNS resident monocytes (microglia) and blood-derived macrophages(Chitnis and Weiner, 2017). Chronic CNS inflammation in the MS lesion is maintained, in part, with pro-inflammatory microglia and macrophages at the rim of chronic active or slowly expanding multiple sclerosis lesions and at the site of ongoing demyelination and axonal damage(Kuhlmann et al., 2017). The continued expansion of chronic active lesions has been postulated to play an essential role in the pathogenesis of disease progression in MS(Frischer et al., 2015).

A significant proportion of microglia and macrophages found at the rim of chronic active MS lesions contain iron, distinguishing them from chronic inactive lesions where iron is essentially absent(Absinta et al., 2016; Bagnato et al., 2011; Dal-Bianco et al., 2017; Mehta et al., 2013). Gradient echo (GRE) MRI sequences are sensitive to iron (Langkammer et al., 2010) and have been used to explore iron dynamics in the brain (Liu et al., 2015). Detection of a paramagnetic rim in a subset of chronic lesions has generated significant interest, and numerous *in vivo* and histopathological validation studies have confirmed the rim is representative of iron-laden inflammatory cells(Absinta et al., 2016; Bagnato et al., 2011; Dal-Bianco et al., 2017; Dal-Bianco et al., 2021). These studies have provided further evidence that chronic active lesions can occur throughout the disease course, including within patients with preclinical disease (Oh et al., 2021; Suthiphosuwan et al., 2020), and can be associated with clinical disability (Absinta et al., 2019). Quantitative susceptibility mapping (QSM) (de Rochefort et al., 2008) is an effective post-processing technique for GRE data to directly map the various sources of susceptibility by solving the field-to-source inversion problem (Yao et al., 2018) and provides a more accurate quantification and localization of brain iron (Deistung et al., 2013; Eskreis-Winkler et al., 2016; Langkammer et al., 2013). Lesions with a hyperintense rim (rim+) on QSM have increased inflammation on PK11195-PET, with histopathological validation confirming a rim of inflammatory cells and more tissue damage on myelin water fraction imaging (Kaunzner et al., 2019; Yao et al., 2018). The quantitative nature of QSM provides a unique opportunity to capture time-dependent changes of rim lesions, as their susceptibility remains high even after years of initial detection and slowly decays over time (Chen et al., 2014; Dal-Bianco et al., 2021; Zhang et al., 2019; Zhang et al., 2016). QSM allows investigators to accurately localize and evaluate inflammatory activity in chronic active lesions, which can provide significant insight into the potential impact of these lesions on disease progression and tissue damage.

The goal of this study was to assess the association of rim+ lesions with clinical disability and global measures of structural tissue damage to further establish QSM as an imaging biomarker for MS. To assess the impact on clinical disability, we explored the relationship between chronic rim+ lesions on both cognitive function and standard measures of neurological disability in a cohort of relapsing remitting (RR) MS patients. Gray matter atrophy, as an MRI measure of neurodegeneration, occurs early and has the strongest association with long-term clinical disability(Azevedo et al., 2018; Eshaghi et al., 2018; Steenwijk et al., 2016); thus, in a secondary analysis, we further explored the impact of rim+ lesions on measures of cortical and thalamic integrity. The ultimate goal of this study was to demonstrate that chronic active MS lesions, as identified by QSM, is associated with the pathogenesis of MS and a potential imaging biomarker to improve disease prognostication.

## Materials and methods

### Patient cohort

This was a cross-sectional study of 159 patients with RRMS meeting the 2010 McDonald criteria(Polman et al., 2011), age ≥ 18 years and who were already participating in our research repository. Patients were recruited to receive a cognitive evaluation and neurological assessment within an average of 14 ± 21 days of their annual MRI scan. Cognitive function was evaluated utilizing *The Brief International Cognitive Assessment for Multiple Sclerosis* (BICAMS), which is composed of three assessments: Symbol Digit Modalities Test (SDMT), California Verbal Learning Test-II (CVLT-II) Immediate Recall (Total of Trials 1-5), and Brief Visuospatial Memory Test-Revised (BVMT-R) Immediate Recall (Total of Trials 1-3)(Langdon et al., 2012). SDMT assesses processing speed, CVLT-II measures verbal learning and short-term memory, and BVMT-R examines visuospatial learning and short-term memory. A lower score for each cognitive assessment is associated with a lower cognitive performance. Expanded Disability Status Score (EDSS) was utilized to measure neurological dysfunction, with a higher EDSS score indicating increased disability. All studies were approved by an ethical standards committee on human experimentation, and written informed consent was obtained from all patients according to the Declaration of Helsinki. The following clinical data was also collected for all patients: gender, age, disease duration from initial symptom, duration on current disease modifying treatments (DMT) and disease subtype.

### MRI protocol and image processing

Imaging was performed on a 3T Magnetom Skyra scanner (Siemens Medical Solutions USA, Malvern, PA) using a product twenty-channel head/neck coil. The typical scanning protocol consisted of sagittal 3D T1-weighted (T1w) MPRAGE sequence (Repetition Time (TR)/Echo Time (TE)/Inversion Time (TI) = 2300/2.3/900 ms, flip angle (FA) = 8°, GRAPPA parallel imaging factor (R) = 2, voxel size = 1.0 × 1.0 × 1.0 mm^3^) prior to Gadolinium administration for anatomical structure and after for detecting blood-brain barrier disruption, axial 2D T2-weighted (T2w) turbo spin echo sequence (TR/TE = 5840/93 ms, FA = 90°, turbo factor = 18, R = 2, voxel size = 0.5 × 0.5 × 3mm^3^) and sagittal 3D fat-saturated T2w fluid attenuated inversion recovery (FLAIR) sequence (TR/TE/TI = 8500/391/2500 ms, FA = 90°, turbo factor = 278, R = 3, voxel size = 1.0 × 1.0 × 1.0 mm^3^) for MS lesion detection, and axial 3D multi-echo GRE sequence for QSM (axial field of view (FOV) = 24 cm, TR/TE1/ΔTE = 48.0/6.3/4.1 ms, number of TEs = 10, FA = 15°, R = 2, voxel size = 0.75×0.93×3 mm^3^, scan time = 4.2 min). QSM was reconstructed from complex GRE images using a fully automated Morphology Enabled Dipole Inversion algorithm zero-referenced to the ventricular cerebrospinal fluid (MEDI+0)(Liu et al., 2018). All the conventional images (T1w, T1w+Gd, T2w, FLAIR) were co-registered to the GRE magnitude images using the FMRIB’s Linear Image Registration Tool algorithm.(Jenkinson et al., 2002)

### Brain tissue segmentation

FreeSurfer(Fischl et al., 1999) was utilized for the segmentation of white matter (WM) and gray matter (GM) on T1w images. The GM segmentation masks were checked and edited for misclassification due to T1-hypointensities associated with lesions.

Cortical thickness measurements were measured from the cortical surface reconstruction results obtained by Freesurfer (Han et al., 2006). Thalamic volumes were obtained using an automated volumetric approach(Fischl et al., 2002), which has been independently validated with respect to the ‘gold standard’ manual segmentation (Keller et al., 2012). Thalamic volume was normalized by overall intracranial volume to account for individual differences in head size.

### Lesion segmentation and QSM rim identification

Multiple sclerosis lesions were segmented on FLAIR images by an automated lesion growth algorithm as implemented in the LST toolbox version 3.0.0 (www.statisticalmodelling.de/lst.html), followed by manual editing and creation of individual lesion labels (Schmidt et al., 2012). Chronic active lesions with paramagnetic rims were identified as by two independent raters, and an additional independent rater then reviewed and resolved the discrepant lesions. As represented in Figure 1, a lesion on FLAIR imaging was labeled as rim+ based on the presence of a hyperintense rim, relative to the lesion core, on QSM imaging. Lesions having a partial or complete rim were considered to be rim+.

**Figure 1:**
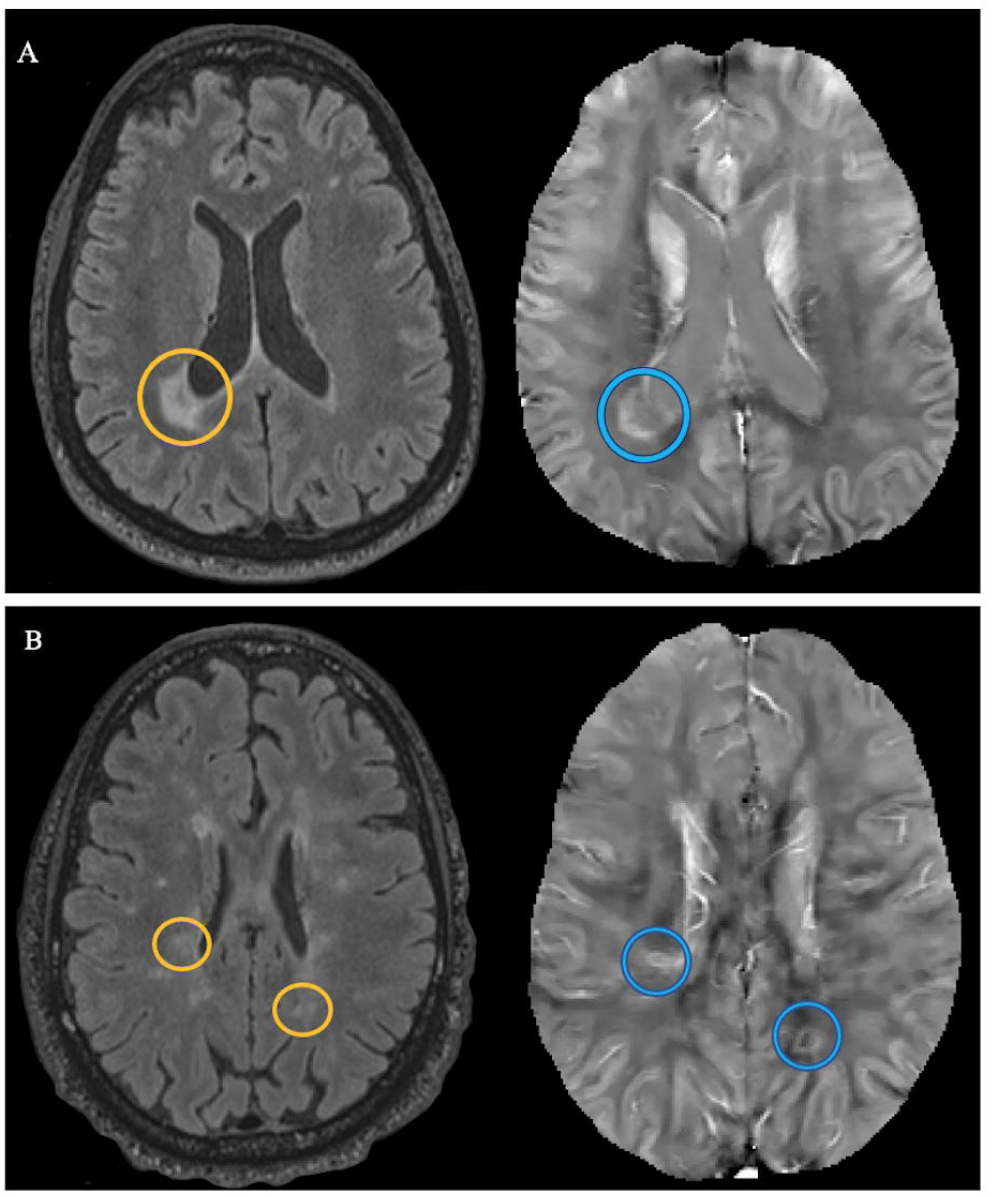
Patient examples of QSM rim+ lesions. Panels A and B represent slices of imaging for two separate patients. Within each panel, FLAIR is located on the left and corresponding slice of QSM is located on the right. In both panels A and B, lesion on FLAIR is located within an orange circle, while corresponding lesion on QSM is located within a blue circle.

The two initial independent raters only disagreed on the classification of 159 lesions yielding an initial agreement of 95.9% (Bangdiwala’s B-statistics= 95.49%). The third reviewer resolved the differences between the two initial raters yielding Cohen’s Kappa coefficients of 70.6% and 85.8% with raters one and two, respectively(Munoz and Bangdiwala, 1997). The Fleiss coefficient for all three raters was equal to 71%.

### Statistical analysis

A multivariate regression model was implemented to test the association between clinical scores (SDMT, CVLT-II, BVMT-R, and EDSS) as a response vector and the number of rim+ lesions per patient, while adjusting for total FLAIR lesion volume (TLV). In addition, the model accounts for other patient level covariates such as gender, age, disease duration and duration of current treatment as well as covariate interactions. The model is multivariate because measures the association between four response variables (SDMT, CVLT-II, BVMT-R, and EDSS) and covariates of interest. A multivariate regression model allows us to quantify the relationship between all four disability scores as response variables and multiple patient level covariates in one framework(Hidalgo and Goodman, 2013), thus providing more powerful tests of significance as compared to running four independent univariate models(Johnson and Wichern, 2007). The number of rim+ lesions per patient had quartiles given by 0,0, and 1 with a maximum value of 17. Based on sample size, data distributions for all variables, and scatterplots with smoothed curves, we concluded that the best definition for the variable number of rim+ lesions is none versus at least one. Model assumptions were checked, and variable transformations were analyzed using descriptive plots and summary tables to improve linearity and normality. All scores (SDMT, CVLT-II, BVMT-R, and EDSS) were analyzed using their original scale. For cognitive measures, we report descriptively the percentage of our sample who fell in the clinically impaired range based on published normative data (Benedict et al., 1996; Delis et al., 2000; Smith, 1991); however, we used raw scores rather than age-normed scores for statistical analyses because age was already included in our models as a covariate. Scatter-plots and summary statistics revealed a linear relationship between all clinical scores and the log-transformed volume (log-TVL).

Two univariate regression models were also implemented to test the association between both cortical thickness and thalamic volume and the number of rim+ lesions at the patient level, while controlling for gender, age, disease duration, duration of current treatment and TLV.

All final models were selected using a stepwise procedure with a 0.10 significance level. Model assumptions were checked. Inferences are based on a 5% significant level. Furthermore model-based means were computed along with their 95% confidence intervals. Statistical analysis was performed using R: A language and environment for statistical computing R Core Team (2020)

## Results

### Patient cohort

This was a cross-sectional study composed of 159 patients with RRMS. The cohort included 115 females and 44 males aged (mean ± SD) 42.17 ± 10.25 years (yrs), current treatment duration of 3.47± 3.97 yrs, disease duration of 10.74 ± 7.51 yrs and median Expanded Disability Status Scale of 1.0 (IQR=2). The proportion of cognitive impairment found among the entire patient cohort was relatively low and observed to be the following: 22.01% on SDMT, 10.06% on CVLT-II, and 25.79% on BVMT-R.

### QSM rim+ lesions

We identified a total of 4,469 lesions, of which 171 (3.8%) classified as rim+ (Fig. 1). The B-test agreement between the two main raters was high (B-statistics= 95.49%). None of the rim+ lesions were enhancing with gadolinium on T1w imaging. The number of QSM rim+ lesions per patient ranged from zero to a maximum of 17 (Median=0, and interquartile range (IQR) =1). One hundred and two patients (64.2%) did not have any rim+ lesions. Of the fifty-seven patients (35.9%) with at least one rim+ lesion, the median number of rim+ lesions per patient was 2 (IQR=3). Table 1 presents the clinical and MRI characteristics among patients without rim+ lesions as compared to those with at least one lesion.

**Table 1.** Clinical and MRI characteristics of patients. Abbreviations include expanded disability status score (EDSS), symbol digit modality test (SDMT), California verbal learning test-II (CVLT-II), and brief visuospatial memory test-revised (BVMT-R). Volumes are in mm^3^, cortical thickness is in mm, and thalamic volume is unitless (normalized to head size). Two-sided p-values were computed using the Welch Two Sample t-test or Mann-Whitney test for continuous symmetric variables. Chi-square test was used to compared categorical variables between the two groups of patients.

| **Characteristics** | **Patients with zero rim+ lesions (n=102)** | **Patients with at least one rim+ lesion (n=57)** | **p-value** |
| --- | --- | --- | --- |
| **Female, n (%)** | 76 (74.51) | 39 (68.42) | 0.523 |
| **Disease Duration, mean (SD)** | 11.02 (8.05) | 10.24 (7.37) | 0.528 |
| **Age, mean (SD)** | 41.96 (17.58) | 42.54 (10.14) | 0.731 |
| **Cognitive Score, mean (SD)** |  |  |  |
| **SDMT** | 58.31 (11.14) | 52.70 (11.78) | **0.004** |
| **CVLT-II** | 54.34 (8.42) | 50.86 (11.95) | **0.028** |
| **BVMT-R** | 25.27 (5.93) | 22.88 (7.57) | 0.032 |
| **Cognitive Impairment, n (%)** |  |  |  |
| **SDMT** | 15 (14.71) | 20 (35.09) | **0.005** |
| **CVLT-II** | 6 (5.88) | 10 (17.54) | **0.038** |
| **BVMT-R** | 21 (20.59) | 20 (35.09) | 0.069 |
| **EDSS, median (IQR )** | 0.5 (2) | 1.50 (2.5) | **0.025** |
| **MRI Measures** |  |  |  |
| **Cortical Thickness, mean (SD)** | 2.53 (0.09) | 2.48 (0.09) | **0.002** |
| **Thalamic Volume, mean (SD)** | 0.0093 (0.0009) | 0.0089 (0.0008) | **0.0051** |
| **No. Lesions on FLAIR, median (IQR)** | 13 (20) | 34 (31) | **<0.001** |
| **Lesion Volume on FLAIR, mean (SD)** | 3127.72 (4619.32) | 10296.04 (10412.46) | **<0.001** |

### Association of QSM rim lesions and disability

Figure 2 compares the sample distributions of cognitive scores of patients with and without rim + lesions. The distributions of SDMT, CVLT-II, BVMT-R have similar spread for both groups of patients, however each sample mean is consistently higher for patients without rim+ lesions.

**Figure 2:**
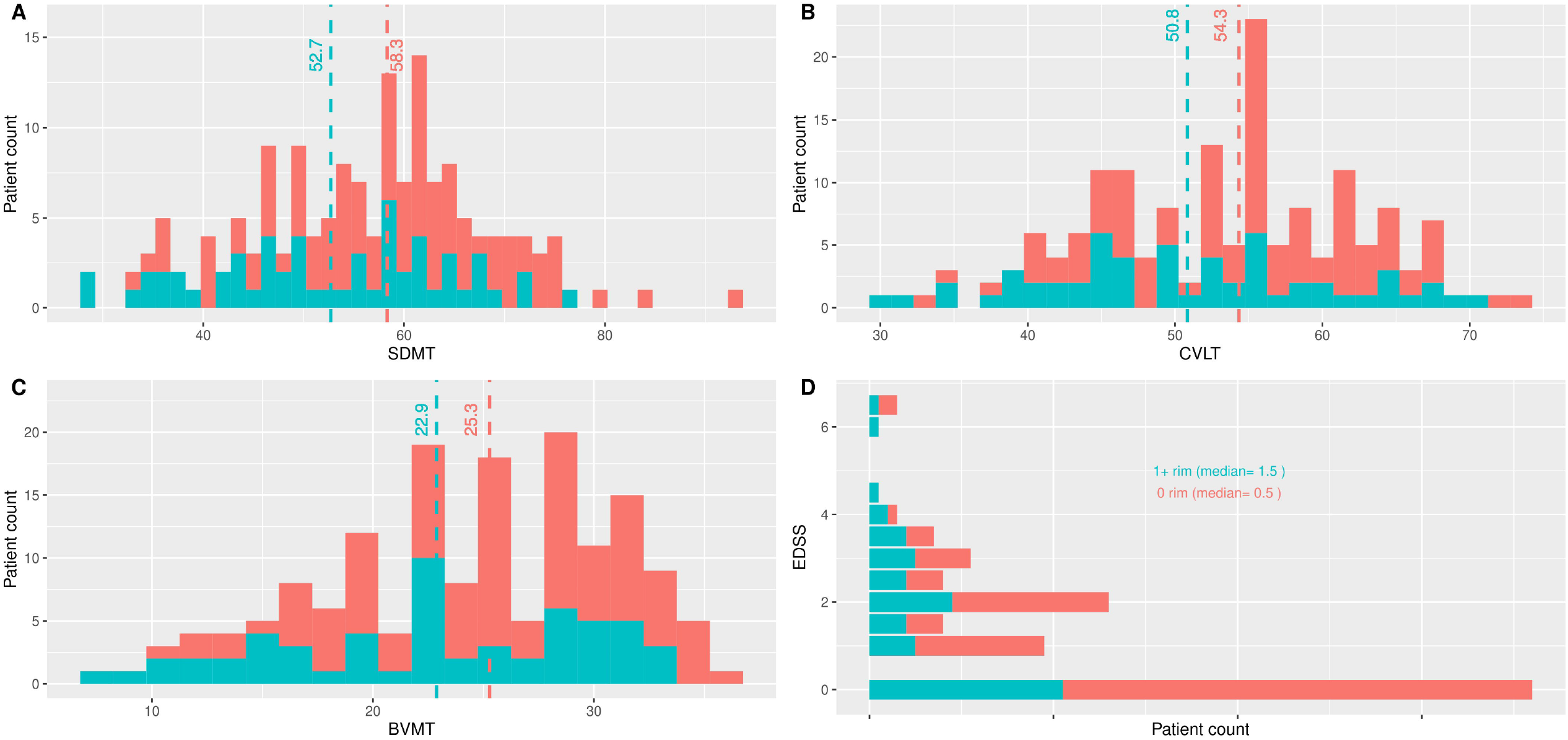
Distribution of disability scores comparing patients with zero (0: red) rim+ lesions versus patients with at least one rim+ lesion (1+: blue). Dash-lines represent mean for each group. Patients with at least one rim+ lesion performed worse on average SDMT [A], CVLT-II [B], BVMT-R [C] and EDSS [D].

Figure 2 also shows that more patients without rim+ lesions have higher SDMT, CVLT-II, BVMT-R scores as the right tails of the plots are scores from exclusively patients without rim+ lesions. The distribution of EDSS also indicates that the majority of patients without rim+ lesions have lower EDSS scores. Given these results, we hypothesized that, on average, patients without rim + lesions have better disability mean scores that patients with at least one rim+ lesion.

A multivariate regression model was used to test our hypothesis and determine if the association between all disability measures (SDMT, CVLT, BVMT-R, and EDSS) and log-transformed total FLAIR lesion volume was dependent on the presence of at least one rim+ lesion after adjusting for other covariates. The final model included treatment duration, age, gender, TLV and the interaction between TLV and at least one rim+ lesion as statistically significant covariates. Table 2 summarizes the p-values obtained from the multivariate analysis of variance (MANOVA) table and the subsequent Analysis of Variance (ANOVA) tables for each clinical score. The final model (MANOVA) included a statistically significant interaction effect of rim+ lesions and total lesion volume on FLAIR imaging on disability outcome measures (p=0.006). Furthermore, number of rim+ lesions (0 versus at least 1) and log-TLV were also significant (p=0.010 and p<0.001, respectively). Other statistically significant patient level covariates were treatment duration, age and gender with p-values of 0.012, <0.001 and 0.038, respectively. The MANOVA p-values above indicate that at least one disability measures (SDMT, CVLT, BVMT-R, and EDSS) is associated with the presence of rim+ lesions.

**Table 2.** Summary of p-values from the Multivariate Analysis of Variance (MANOVA) table and subsequent Analysis of Variance (ANOVA) tables for each score. The final multivariate model for SDMT, CVLT-II, BVMT-R, and EDSS as vector of response variables included current Treatment Duration, Age, Gender, No. RIM of+ lesions, log T2wFLAIR lesion volume, and the interaction term No. RIM * logT2wFLAIR.lesion.volume as covariates. This report gives the p-values associated with the MANOVA Pillai test and the approximated F-statistics for each score (ANOVA)

|  | MANOVA | ANOVA for each Cognitive Score |  |  |  |
| --- | --- | --- | --- | --- | --- |
|  |  | SDMT | CVLT-II | BVMT-R | EDSS |
| **(Intercept)** | <0.001 | <0.001 | <0.001 | <0.001 | 0.386 |
| **Current Treatment Duration** | 0.012 | 0.389 | 0.137 | 0.513 | 0.009 |
| **Age** | <0.001 | <0.001 | 0.030 | <0.001 | <0.001 |
| **Gender** | 0.038 | 0.508 | 0.003 | 0.592 | 0.841 |
| **No. RIM (0 rim+ versus 1+ rim+)** | 0.010 | 0.011 | 0.050 | 0.157 | 0.001 |
| **logT2wFLAIR.lesion.volume** | <0.001 | <0.001 | 0.294 | 0.002 | 0.607 |
| **No. RIM \***<br>**logT2wFLAIR.lesion.volume** | 0.006 | 0.010 | 0.030 | 0.165 | 0.001 |

Individual analysis of variance tables for each score confirm that the interaction effect between log-TLV and number of rim+ lesions (0 versus at least 1) was statistically significant for SDMT 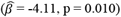, CVLT-II 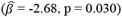, and EDSS 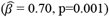. That is, as overall total lesion burden increases, patients with at least one rim+ lesion on quantitative susceptibility mapping have, on average, lower SDMT and CVLT-II mean scores. As TLV increases, EDSS mean scores for patients with at least one rim+ lesions are higher than for patients without rim+ lesions

Figure 3 displays a graphical representation of model based cognitive and EDSS mean difference estimates between patients with zero rim+ lesions versus at least one rim+ lesion and their 95% confidence intervals. These results highlight the impact the interaction that exists with both increasing TLV and the presence of only one rim+ lesion. As TLV increases, the difference between mean SDMT (and CVLT-II) scores between patients without rim+ and at least one rim+ becomes significant and shown with 95% confidence intervals that do not include zero. This implies that patients without rim+ have statistically significant higher mean SDMT and CVLT-II scores than patients with at least one rim+ lesion as TVL increases. Similarly, mean EDSS increased faster for patients with at least one rim+ lesion. There was no significant difference in performance found between groups for BVMT-R.

**Figure 3:**
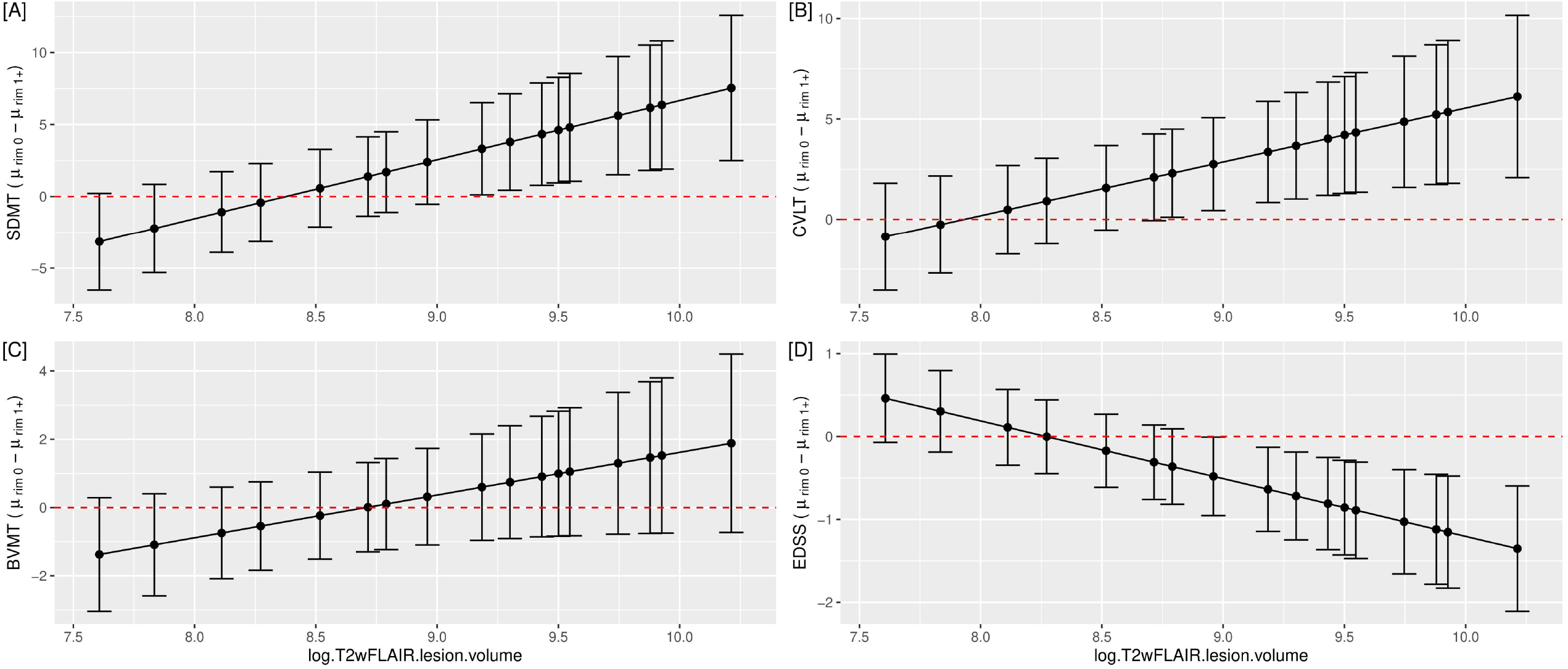
Association of rim+ lesions with disability measures after accounting for clinical and imaging covariates. Patients with both higher total lesion volume on FLAIR and at least one rim+ lesion performed worse on SDMT [A], CVLT-II [B], and EDSS [D] as demonstrated by 95% confidence intervals of the difference of mean disability scores between patients with zero rim+ minus patients with at least one rim+ lesion. The 95% confidence intervals of the means are model-based estimates. The final model included treatment duration, age, gender, total lesion volume on FLAIR, and number of rim+ (statistically significant covariates).

### Association of QSM rim lesions and gray matter damage

In this secondary analysis, we modeled the association between cortical thinning and having a rim+ lesion 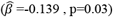 while controlling for other covariates. The final model also included age 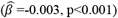 importantly, TLV was not found to be associated with cortical thickness p>0.10). Figure 4 demonstrates the results of the model. The separation of confidence intervals shows the impact of rim+ lesions on cortical thickness, wherein younger-aged patients, with at least one rim+ lesion, have significantly more cortical thinning as compared to patients without a rim+ lesion. As patients age beyond 45 years, aging confounds the influence of a rim+ lesion and eventually becomes less relevant. For both patient cohorts, cortical thickness decreased with increasing age. In a separate model, rim+ lesions were not statistically significant associated with lower thalamic volume (p>0.10), after controlling for age 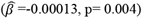, gender 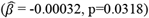, and TLV 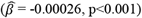,

**Figure 4:**
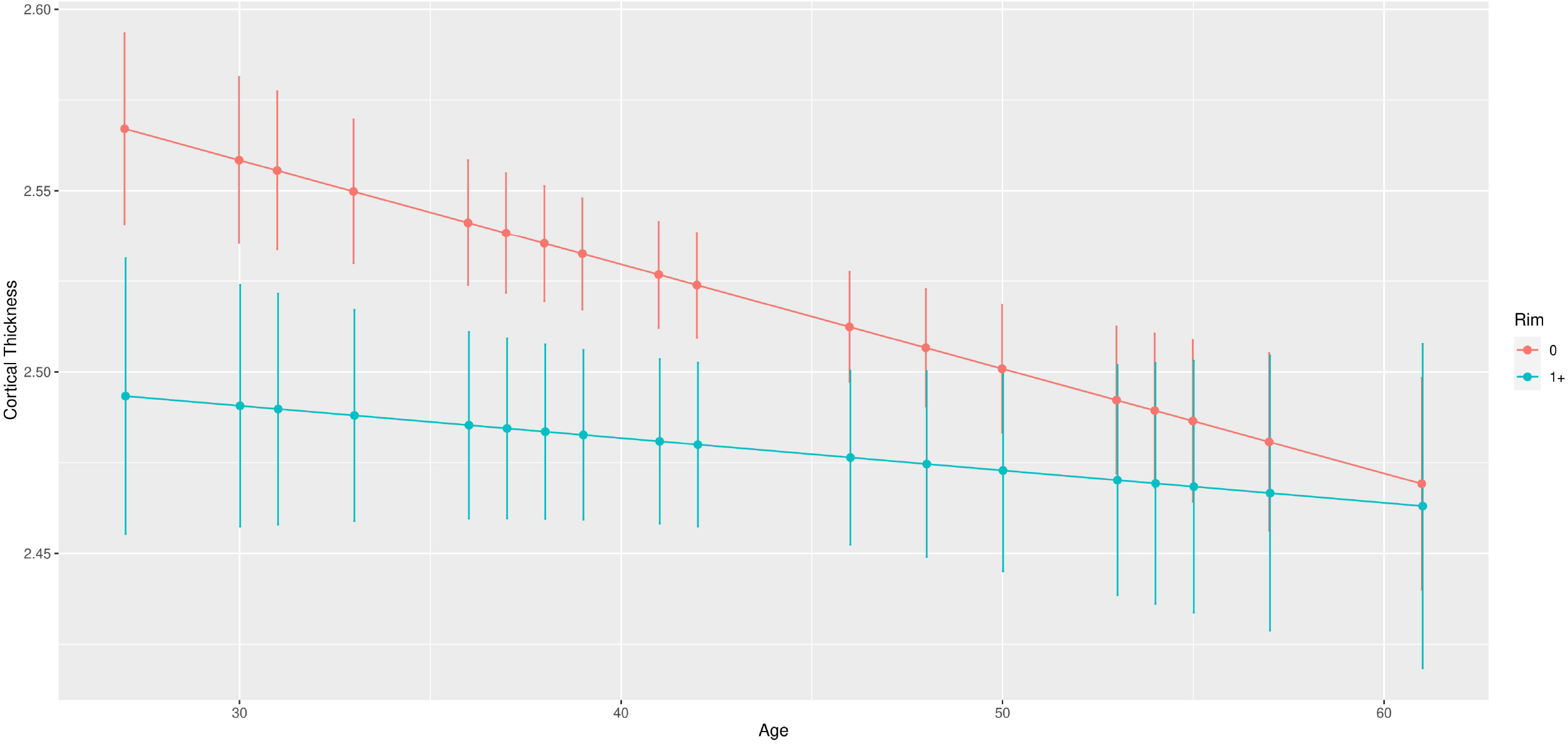
Association between cortical thickness (CT) and age classified by rim+ lesions. 95% confidence intervals for model-based means are represented after adjusting for patient covariates. The final model included age and number of rim+ (statistically significant covariates).

## Discussion

In this study, we utilized QSM to assess the association of chronic active MS lesions with clinical disability, including cognition, and gray matter damage, in a large cohort of patients with multiple sclerosis. We were able to demonstrate the meaningful impact of having at least one QSM rim+ lesion on both the clinical and imaging outcomes.

The identification of rim+ lesions using QSM is based upon the visualization of a hyperintense signal, reflective of increased susceptibility due to iron deposition, at the rim of a subset of chronic lesions (Wisnieff et al., 2014). QSM provides a method to processes GRE phase data (Dong et al., 2015) and deconvolute the magnetic field to uncover the magnetic properties of tissues, localize the tissue magnetic source, and quantify tissue susceptibility; as a result, it provides an accurate localization of iron deposition (Stuber et al., 2016; Wang and Liu, 2015; Wang et al., 2017). Our study harnessed the capacity of QSM to successfully identify lesions with a rim of iron-laden microglia and macrophages (Kaunzner et al., 2019; Yao et al., 2018) and assess their relationship with disability in patients with multiple sclerosis.

A multivariate model was selected to assess the relationship between rim+ lesions and disability given that this approach can account for the correlation between multivariate clinical scores for an individual patients’ covariates. We found a significant joint effect of TLV the number of rim+ lesions on clinical scores. This observation highlights the importance of appropriately considering the combined joint effect of chronic active lesions and total lesions on clinical outcomes. In this study, we demonstrated that in patients with at least one rim+ lesion, as the overall FLAIR lesion burden increases, both cognition and EDSS worsen, reflective of the linear relationship of the two imaging variables. The relationship between chronic active MS lesions and performance on EDSS and processing speed, as measured by the SDMT, has been previously reported (Absinta et al., 2019); however, this study has essential differences. Our work expands upon these recent findings by incorporating the relationship with overall lesion volume and expanding to include additional domains of cognition, including verbal and visuospatial memory as measured by the CVLT-II and BVMT-R, respectively. Through the use of QSM, we were able to identify the impact of one or more chronic active multiple sclerosis lesions on disability, while in the study of Absinta et al. the impact of phase rim lesions on disability was only associated with four or more phase rim lesions(Absinta et al., 2019). We speculate this difference is related to technical differences(Cronin et al., 2016; Eskreis-Winkler et al., 2016) and warrants a thorough evaluation to determine which imaging approach most accurately identifies this subset of chronic lesions.

Importantly, a minority of our patients would be classified as cognitively impaired, and given this, as well as the fact that age was already accounted for in statistical models, we chose to keep the raw cognitive scores for the analysis. Our findings suggest that the relationship between total lesion burden, chronic active lesions, and cognition, which may be especially relevant for early, pre-clinical cognitive changes in MS. Such subtle, early decline in processing speed and memory is functionally impactful, particularly for young patients who are likely to have significant social and occupational responsibilities. Our results may be informative in identifying patients that might benefit the most from early cognitive interventions. Although no significant results were found regarding the relationship between rim+ lesions and BVMT-R, a decreased performance was found as TLV increased. Attributing cognitive dysfunction to white matter lesion characteristics, mainly lesion volume, has yielded variable results in past studies (Rocca et al., 2015), likely related to the dominant role of cortical pathology (Louapre et al., 2016) or the lack of consideration of the white matter lesions’ impact on the brain’s overall connectivity network. In fact, our recent study revealed that, while cortical/subcortical pathology indeed was strongly predictive of cognitive outcomes, adding information about the topological impact of white matter lesions on the brain’s connectivity network may improve prediction accuracy (Kuceyeski et al., 2018). Therefore, even our positive results regarding SDMT and CVLT-II should be interpreted with caution. In addition, *BICAMS* is a limited battery of cognitive assessments and not meant to be a comprehensive neuropsychological evaluation (Langdon et al., 2012).

In a secondary analysis, we explored the influence of having at least one rim+ lesion on measures of gray matter integrity, with a goal to demonstrate an association of chronic active multiple sclerosis lesions with global neurodegeneration. A lower thalamic volume has been previously associated with having higher numbers of chronic active lesions, as measured by phase imaging (Absinta et al., 2019), although patients with QSM rim lesions had a smaller thalamic volume in the current study, the difference failed to reach significance in the regression model. Our results identified a strong influence of total lesion burden on thalamic volume, suggesting that diffuse damage to white matter connections may have a stronger influence on thalamic damage(Cifelli et al., 2002). Nevertheless, our study is unique in showing the strong association with cortical gray matter. Interestingly, overall lesion volume was not associated with this outcome, but as expected, age was a significant covariate, and its influence on cortical gray matter dominated over the effect of having a rim+ lesion at older ages. These results highlight the importance of chronic active lesions on the overall neurodegenerative process in MS and, unique to this study, demonstrate an impact at earlier stages of the disease. It can be speculated that the slow expansion of these lesions, with ongoing demyelination and axonal loss (Kuhlmann et al., 2017) and increased oxidative stress from the presence of activated microglia (Lassmann, 2018), contribute more to these observed global degenerative changes. As larger studies explore the influence of rim+ lesions on disease course, further validation of these results is warranted, but our results do suggest that early identification of rim+ lesions is a potential treatment target to influence the ongoing and subsequent neurodegenerative process in MS.

Chronic active MS lesions have traditionally been associated with progressive disease, which is based upon post-mortem histopathological studies (Frischer et al., 2015). However, given the bias toward progressive patients within these studies, it is unclear how prevalent chronic active lesions are in earlier stages of the disease. Studies utilizing phase imaging have demonstrated that over 50% of patients have at least one phase rim+ lesion in relapsing disease, and phase rim+ lesions, as consistent with post-mortem studies, appear to be more common in those with progressive disease (Absinta et al., 2019; Maggi et al., 2020). Interestingly, it was recently demonstrated that, at the preclinical stage of the disease, or with radiologic isolated syndrome, phase rim lesions occurred in 61% of patients and included 12% of overall lesions (Suthiphosuwan et al., 2020), whereas chronic lesions in patients with multiple sclerosis are reported to represent approximately one-third of the lesions (Absinta et al., 2018). In our study, rim+ lesions represent a minority of chronic lesions (3.8%), and only 35.9% of patients had a rim+ lesion; again, this discrepancy may relate to differences between phase and QSM. Although other studies have similarly found that QSM rim+ lesions represent a minority of white matter lesions (10-20%)(Harrison et al., 2016; Kaunzner et al., 2019; Yao et al., 2018), the amount of rim+ lesions in this patient cohort is relatively low. The variability in these results may be related to differences among the cohorts and lack of a specific criterion for identifying a hyperintense rim. Similar to other paramagnetic rim studies, lesions are characterized visually to define a QSM rim+ lesion and variability in this definition among groups of readers is likely a contributing factor. Reviewers for this study were clearly conservative in their definition but importantly, demonstrated a strong agreement. Our current slice thickness of 3 mm could limit our detection of rims; however, our sequence provides high in-plane resolution for improved excellent axial visualization and can be acquired within 4 minutes, making this a clinically feasible sequence. As we move forward, larger, multi-center studies should standardize the most accurate imaging approach and rim lesion definition. Regardless of our low rim+ lesion number, our results still indicated that the presence of these lesions was associated with lower scores on disability and more cortical gray matter loss.

There were limitations to this study. The clinical cohort consisted of RRMS patients with a relatively low disease duration and disease burden. To obtain a more comprehensive picture of the impact of QSM rim+ lesions on cognition and physical disability, it is possible that a more impaired population, such as one with a large representation of progressive patients, may be more informative. Furthermore, our study was observational and only association statements can be drawn.

In conclusion, our study demonstrates the significant impact of chronic active lesions on both clinical and imaging features of disability and highlights the relevance of identifying these lesions at each stage of the disease. Furthermore, our work promotes the use of QSM to identify chronic active lesions and as a tool to understand mechanisms of injury leading to clinical progression and neurodegeneration. Future work will include longitudinal studies building upon our findings to establish the role of rim+ lesions as a prognostic biomarker in MS and to further explore the quantitative aspects of QSM to assess response to novel therapeutic approaches targeting the innate immune response in the CNS.

## Data Availability

The data that support the findings of this study are available from the corresponding author upon reasonable request.

## Acknowledgements

The research conducted in this study would not be possible without the patients at the Judith Jaffe Multiple Sclerosis Center. We would also like to acknowledge Sneha Pandya and Dr. Weiyuan Huang for their advisement with the analysis and Dr. Timothy Vartanian for his review of the manuscript. This study was supported in part by grants R01NS104283, 090464, 105144 and R21NS104634 from the National Institutes of Health and by grant UL1 TR000456-06 from the Weill Cornell Clinical and Translational Science Center (CTSC). Sponsors did not have a role in the study design, in the collection, analysis, interpretation of data, in the writing of the report or in the decision to submit the article for publication.

## Abbreviations

(MS): multiple sclerosis
(RR): relapsing remitting
(BVMT-R): Brief Visuospatial Memory Test-Revised
(CVLT-II): California Verbal Learning Test-II
(EDSS): expanded disability status score
(FLAIR): fluid attenuated inversion recovery imaging
(QSM): quantitative susceptibility mapping
(SDMT): Symbol Digit Modalities Test

## References

Absinta, M., Sati, P., Fechner, A., Schindler, M.K., Nair, G., Reich, D.S., 2018. Identification of Chronic Active Multiple Sclerosis Lesions on 3T MRI. AJNR Am J Neuroradiol 39, 1233–1238.

Absinta, M., Sati, P., Masuzzo, F., Nair, G., Sethi, V., Kolb, H., Ohayon, J., Wu, T., Cortese, I.C.M., Reich, D.S., 2019. Association of Chronic Active Multiple Sclerosis Lesions With Disability In Vivo. JAMA Neurol.

Absinta, M., Sati, P., Schindler, M., Leibovitch, E.C., Ohayon, J., Wu, T., Meani, A., Filippi, M., Jacobson, S., Cortese, I.C., Reich, D.S., 2016. Persistent 7-tesla phase rim predicts poor outcome in new multiple sclerosis patient lesions. J Clin Invest.

Azevedo, C.J., Cen, S.Y., Khadka, S., Liu, S., Kornak, J., Shi, Y., Zheng, L., Hauser, S.L., Pelletier, D., 2018. Thalamic atrophy in multiple sclerosis: A magnetic resonance imaging marker of neurodegeneration throughout disease. Ann Neurol 83, 223–234.

Bagnato, F., Hametner, S., Yao, B., van Gelderen, P., Merkle, H., Cantor, F.K., Lassmann, H., Duyn, J.H., 2011. Tracking iron in multiple sclerosis: a combined imaging and histopathological study at 7 Tesla. Brain 134, 3602–3615.

Benedict, R.H.B., Schretlen, D., Groninger, L., Dobraski, M., Shpritz, B., 1996. Revision of the brief visuospatial memory test: Studies of normal performance, reliability, and validity. Psychological Assessment 8, 145–153.

Chen, W., Gauthier, S.A., Gupta, A., Comunale, J., Liu, T., Wang, S., Pei, M., Pitt, D., Wang, Y., 2014. Quantitative susceptibility mapping of multiple sclerosis lesions at various ages. Radiology 271, 183–192.

Chitnis, T., Weiner, H.L., 2017. CNS inflammation and neurodegeneration. J Clin Invest 127, 3577–3587.

Cifelli, A., Arridge, M., Jezzard, P., Esiri, M.M., Palace, J., Matthews, P.M., 2002. Thalamic neurodegeneration in multiple sclerosis. Ann Neurol 52, 650–653.

Cronin, M.J., Wharton, S., Al-Radaideh, A., Constantinescu, C., Evangelou, N., Bowtell, R., Gowland, P.A., 2016. A comparison of phase imaging and quantitative susceptibility mapping in the imaging of multiple sclerosis lesions at ultrahigh field. MAGMA 29, 543–557.

Dal-Bianco, A., Grabner, G., Kronnerwetter, C., Weber, M., Hoftberger, R., Berger, T., Auff, E., Leutmezer, F., Trattnig, S., Lassmann, H., Bagnato, F., Hametner, S., 2017. Slow expansion of multiple sclerosis iron rim lesions: pathology and 7 T magnetic resonance imaging. Acta Neuropathol 133, 25–42.

Dal-Bianco, A., Grabner, G., Kronnerwetter, C., Weber, M., Kornek, B., Kasprian, G., Berger, T., Leutmezer, F., Rommer, P.S., Trattnig, S., Lassmann, H., Hametner, S., 2021. Long-term evolution of multiple sclerosis iron rim lesions in 7 T MRI. Brain.

de Rochefort, L., Brown, R., Prince, M.R., Wang, Y., 2008. Quantitative MR susceptibility mapping using piece-wise constant regularized inversion of the magnetic field. Magn Reson Med 60, 1003–1009.

Deistung, A., Schafer, A., Schweser, F., Biedermann, U., Turner, R., Reichenbach, J.R., 2013. Toward in vivo histology: a comparison of quantitative susceptibility mapping (QSM) with magnitude-, phase-, and R2*-imaging at ultra-high magnetic field strength. Neuroimage 65, 299– 314.

Delis, D., Kramer, J., Kaplan, E., Ober, B., 2000. California Verbal Learning Test Second Edition Adult version. Manual, second ed. Pearson, Bloomington, MN.

Dong, J., Liu, T., Chen, F., Zhou, D., Dimov, A., Raj, A., Cheng, Q., Spincemaille, P., Wang, Y., 2015. Simultaneous phase unwrapping and removal of chemical shift (SPURS) using graph cuts: application in quantitative susceptibility mapping. IEEE Trans Med Imaging 34, 531–540.

Eshaghi, A., Marinescu, R.V., Young, A.L., Firth, N.C., Prados, F., Jorge Cardoso, M., Tur, C., De Angelis, F., Cawley, N., Brownlee, W.J., De Stefano, N., Laura Stromillo, M., Battaglini, M., Ruggieri, S., Gasperini, C., Filippi, M., Rocca, M.A., Rovira, A., Sastre-Garriga, J., Geurts, J.J.G., Vrenken, H., Wottschel, V., Leurs, C.E., Uitdehaag, B., Pirpamer, L., Enzinger, C., Ourselin, S., Gandini Wheeler-Kingshott, C.A., Chard, D., Thompson, A.J., Barkhof, F., Alexander, D.C., Ciccarelli, O., 2018. Progression of regional grey matter atrophy in multiple sclerosis. Brain 141, 1665–1677.

Eskreis-Winkler, S., Zhou, D., Liu, T., Gupta, A., Gauthier, S.A., Wang, Y., Spincemaille, P., 2016. On the influence of zero-padding on the nonlinear operations in Quantitative Susceptibility Mapping. Magn Reson Imaging 35, 154–159.

Fischl, B., Salat, D.H., Busa, E., Albert, M., Dieterich, M., Haselgrove, C., van der Kouwe, A., Killiany, R., Kennedy, D., Klaveness, S., Montillo, A., Makris, N., Rosen, B., Dale, A.M., 2002. Whole brain segmentation: automated labeling of neuroanatomical structures in the human brain. Neuron 33, 341–355.

Fischl, B., Sereno, M.I., Dale, A.M., 1999. Cortical surface-based analysis. II: Inflation, flattening, and a surface-based coordinate system. Neuroimage 9, 195–207.

Frischer, J.M., Weigand, S.D., Guo, Y., Kale, N., Parisi, J.E., Pirko, I., Mandrekar, J., Bramow, S., Metz, I., Bruck, W., Lassmann, H., Lucchinetti, C.F., 2015. Clinical and pathological insights into the dynamic nature of the white matter multiple sclerosis plaque. Ann Neurol 78, 710–721.

Han, X., Jovicich, J., Salat, D., van der Kouwe, A., Quinn, B., Czanner, S., Busa, E., Pacheco, J., Albert, M., Killiany, R., Maguire, P., Rosas, D., Makris, N., Dale, A., Dickerson, B., Fischl, B., 2006. Reliability of MRI-derived measurements of human cerebral cortical thickness: the effects of field strength, scanner upgrade and manufacturer. Neuroimage 32, 180–194.

Harrison, D.M., Li, X., Liu, H., Jones, C.K., Caffo, B., Calabresi, P.A., van Zijl, P., 2016. Lesion Heterogeneity on High-Field Susceptibility MRI Is Associated with Multiple Sclerosis Severity. AJNR Am J Neuroradiol 37, 1447–1453.

Hidalgo, B., Goodman, M., 2013. Multivariate or multivariable regression? Am J Public Health 103, 39–40.

Jenkinson, M., Bannister, P., Brady, M., Smith, S., 2002. Improved optimization for the robust and accurate linear registration and motion correction of brain images. Neuroimage 17, 825–841.

Johnson, R.A., Wichern, D.W., 2007. Applied Multivariate Statistical Analysis,. Pearson Prentice-Hall., Upper Saddle River, NJ.

Kaunzner, U.W., Kang, Y., Zhang, S., Morris, E., Yao, Y., Pandya, S., Hurtado Rua, S.M., Park, C., Gillen, K.M., Nguyen, T.D., Wang, Y., Pitt, D., Gauthier, S.A., 2019. Quantitative susceptibility mapping identifies inflammation in a subset of chronic multiple sclerosis lesions. Brain 142, 133–145.

Keller, S.S., Gerdes, J.S., Mohammadi, S., Kellinghaus, C., Kugel, H., Deppe, K., Ringelstein, E.B., Evers, S., Schwindt, W., Deppe, M., 2012. Volume estimation of the thalamus using freesurfer and stereology: consistency between methods. Neuroinformatics 10, 341–350.

Kuceyeski, A., Monohan, E., Morris, E., Fujimoto, K., Vargas, W., Gauthier, S.A., 2018. Baseline biomarkers of connectome disruption and atrophy predict future processing speed in early multiple sclerosis. Neuroimage Clin 19, 417–424.

Kuhlmann, T., Ludwin, S., Prat, A., Antel, J., Bruck, W., Lassmann, H., 2017. An updated histological classification system for multiple sclerosis lesions. Acta Neuropathol 133, 13–24.

Langdon, D.W., Amato, M.P., Boringa, J., Brochet, B., Foley, F., Fredrikson, S., Hamalainen, P., Hartung, H.P., Krupp, L., Penner, I.K., Reder, A.T., Benedict, R.H., 2012. Recommendations for a Brief International Cognitive Assessment for Multiple Sclerosis (BICAMS). Mult Scler 18, 891–898.

Langkammer, C., Krebs, N., Goessler, W., Scheurer, E., Ebner, F., Yen, K., Fazekas, F., Ropele, S., 2010. Quantitative MR imaging of brain iron: a postmortem validation study. Radiology 257, 455–462.

Langkammer, C., Liu, T., Khalil, M., Enzinger, C., Jehna, M., Fuchs, S., Fazekas, F., Wang, Y., Ropele, S., 2013. Quantitative susceptibility mapping in multiple sclerosis. Radiology 267, 551– 559.

Lassmann, H., 2018. Multiple Sclerosis Pathology. Cold Spring Harb Perspect Med 8.

Liu, C., Li, W., Tong, K.A., Yeom, K.W., Kuzminski, S., 2015. Susceptibility-weighted imaging and quantitative susceptibility mapping in the brain. J Magn Reson Imaging 42, 23–41.

Liu, Z., Spincemaille, P., Yao, Y., Zhang, Y., Wang, Y., 2018. MEDI+0: Morphology enabled dipole inversion with automatic uniform cerebrospinal fluid zero reference for quantitative susceptibility mapping. Magn Reson Med 79, 2795–2803.

Louapre, C., Govindarajan, S.T., Gianni, C., Cohen-Adad, J., Gregory, M.D., Nielsen, A.S., Madigan, N., Sloane, J.A., Kinkel, R.P., Mainero, C., 2016. Is the Relationship between Cortical and White Matter Pathologic Changes in Multiple Sclerosis Spatially Specific? A Multimodal 7- T and 3-T MR Imaging Study with Surface and Tract-based Analysis. Radiology 278, 524–535.

Maggi, P., Sati, P., Nair, G., Cortese, I.C.M., Jacobson, S., Smith, B.R., Nath, A., Ohayon, J., van Pesch, V., Perrotta, G., Pot, C., Theaudin, M., Martinelli, V., Scotti, R., Wu, T., Du Pasquier, R., Calabresi, P.A., Filippi, M., Reich, D.S., Absinta, M., 2020. Paramagnetic Rim Lesions Are Specific to Multiple Sclerosis: An International Multicenter 3t Mri Study. Ann Neurol.

Mehta, V., Pei, W., Yang, G., Li, S., Swamy, E., Boster, A., Schmalbrock, P., Pitt, D., 2013. Iron is a sensitive biomarker for inflammation in multiple sclerosis lesions. PLoS One 8, e57573.

Munoz, S.R., Bangdiwala, S.I., 1997. Interpretation of Kappa and B statistics measures of agreement. Journal of Applied Statistics 24, 105–111.

Oh, J., Suthiphosuwan, S., Sati, P., Absinta, M., Dewey, B., Guenette, M., Selchen, D., Bharatha, A., Donaldson, E., Reich, D.S., Feinstein, A., 2021. Cognitive impairment, the central vein sign, and paramagnetic rim lesions in RIS. Mult Scler, 13524585211002097.

Polman, C.H., Reingold, S.C., Banwell, B., Clanet, M., Cohen, J.A., Filippi, M., Fujihara, K., Havrdova, E., Hutchinson, M., Kappos, L., Lublin, F.D., Montalban, X., O’Connor, P., Sandberg-Wollheim, M., Thompson, A.J., Waubant, E., Weinshenker, B., Wolinsky, J.S., 2011. Diagnostic criteria for multiple sclerosis: 2010 revisions to the McDonald criteria. Ann Neurol 69, 292–302.

Rocca, M.A., Amato, M.P., De Stefano, N., Enzinger, C., Geurts, J.J., Penner, I.K., Rovira, A., Sumowski, J.F., Valsasina, P., Filippi, M., Group, M.S., 2015. Clinical and imaging assessment of cognitive dysfunction in multiple sclerosis. Lancet Neurol 14, 302–317.

Schmidt, P., Gaser, C., Arsic, M., Buck, D., Forschler, A., Berthele, A., Hoshi, M., Ilg, R., Schmid, V.J., Zimmer, C., Hemmer, B., Muhlau, M., 2012. An automated tool for detection of FLAIR-hyperintense white-matter lesions in Multiple Sclerosis. Neuroimage 59, 3774–3783.

Smith, A., 1991. Symbol digit modalities test: Manual.. Western Psychological Services, Los Angeles, CA.

Steenwijk, M.D., Geurts, J.J., Daams, M., Tijms, B.M., Wink, A.M., Balk, L.J., Tewarie, P.K., Uitdehaag, B.M., Barkhof, F., Vrenken, H., Pouwels, P.J., 2016. Cortical atrophy patterns in multiple sclerosis are non-random and clinically relevant. Brain 139, 115–126.

Stuber, C., Pitt, D., Wang, Y., 2016. Iron in Multiple Sclerosis and Its Noninvasive Imaging with Quantitative Susceptibility Mapping. Int J Mol Sci 17.

Suthiphosuwan, S., Sati, P., Absinta, M., Guenette, M., Reich, D.S., Bharatha, A., Oh, J., 2020. Paramagnetic Rim Sign in Radiologically Isolated Syndrome. JAMA Neurol 77, 653–655.

Wang, Y., Liu, T., 2015. Quantitative susceptibility mapping (QSM): decoding MRI data for a tissue magnetic biomarker. Magnetic Resonance in Medicine 73, 82–101.

Wang, Y., Spincemaille, P., Liu, Z., Dimov, A., Deh, K., Li, J., Zhang, Y., Yao, Y., Gillen, K.M., Wilman, A.H., Gupta, A., Tsiouris, A.J., Kovanlikaya, I., Chiang, G.C., Weinsaft, J.W., Tanenbaum, L., Chen, W., Zhu, W., Chang, S., Lou, M., Kopell, B.H., Kaplitt, M.G., Devos, D., Hirai, T., Huang, X., Korogi, Y., Shtilbans, A., Jahng, G.H., Pelletier, D., Gauthier, S.A., Pitt, D., Bush, A.I., Brittenham, G.M., Prince, M.R., 2017. Clinical quantitative susceptibility mapping (QSM): Biometal imaging and its emerging roles in patient care. J Magn Reson Imaging 46, 951–971.

Wisnieff, C., Ramanan, S., Olesik, J., Gauthier, S., Wang, Y., Pitt, D., 2014. Quantitative susceptibility mapping (QSM) of white matter multiple sclerosis lesions: Interpreting positive susceptibility and the presence of iron. Magn Reson Med.

Yao, Y., Nguyen, T.D., Pandya, S., Zhang, Y., Hurtado Rua, S., Kovanlikaya, I., Kuceyeski, A., Liu, Z., Wang, Y., Gauthier, S.A., 2018. Combining Quantitative Susceptibility Mapping with Automatic Zero Reference (QSM0) and Myelin Water Fraction Imaging to Quantify Iron-Related Myelin Damage in Chronic Active MS Lesions. AJNR Am J Neuroradiol 39, 303–310.

Zhang, S., Nguyen, T.D., Hurtado Rua, S.M., Kaunzner, U.W., Pandya, S., Kovanlikaya, I., Spincemaille, P., Wang, Y., Gauthier, S.A., 2019. Quantitative Susceptibility Mapping of Time-Dependent Susceptibility Changes in Multiple Sclerosis Lesions. AJNR Am J Neuroradiol.

Zhang, Y., Gauthier, S.A., Gupta, A., Comunale, J., Chia-Yi Chiang, G., Zhou, D., Chen, W., Giambrone, A.E., Zhu, W., Wang, Y., 2016. Longitudinal change in magnetic susceptibility of new enhanced multiple sclerosis (MS) lesions measured on serial quantitative susceptibility mapping (QSM). J Magn Reson Imaging.

